# Understanding the Potential Impact of Different Drug Properties On SARS-CoV-2 Transmission and Disease Burden: A Modelling Analysis

**DOI:** 10.1101/2021.06.17.21259078

**Authors:** Charles Whittaker, Oliver J. Watson, Carlos Alvarez-Moreno, Nasikarn Angkasekwinai, Adhiratha Boonyasiri, Luis Carlos Triana, Duncan Chanda, Lantharita Charoenpong, Methee Chayakulkeeree, Graham S. Cooke, Julio Croda, Zulma M Cucunubá, Bimandra A. Djaafara, Cassia F. Estofolete, Maria Eugenia Grillet, Nuno R. Faria, Silvia Figueiredo Costa, David A. Forero-Peña, Diana M. Gibb, Anthony C Gordon, Raph L. Hamers, Arran Hamlet, Vera Irawany, Anupop Jitmuang, Nukool Keurueangkul, Teresia Njoki Kimani, Margarita Lampo, Anna S. Levin, Gustavo Lopardo, Rima Mustafa, Shevanthi Nayagam, Thundon Ngamprasertchai, Ng’ang’a Irene Hannah Njeri, Mauricio L. Nogueira, Esteban Ortiz-Prado, Mauricio W. Perroud, Andrew N. Phillips, Panuwat Promsin, Ambar Qavi, Alison J. Rodger, Ester C. Sabino, Sorawat Sangkaew, Djayanti Sari, Rujipas Sirijatuphat, Andrei C. Sposito, Pratthana Srisangthong, Hayley A. Thompson, Zarir Udwadia, Sandra Valderrama-Beltrán, Peter Winskill, Azra C. Ghani, Patrick G.T. Walker, Timothy B. Hallett

## Abstract

**Background:** The unprecedented public health impact of the COVID-19 pandemic has motivated a rapid search for potential therapeutics, with some key successes. However, the potential impact of different treatments, and consequently research and procurement priorities, have not been clear.

**Methods and Findings:** We develop a mathematical model of SARS-CoV-2 transmission, COVID-19 disease and clinical care to explore the potential public-health impact of a range of different potential therapeutics, under a range of different scenarios varying: i) healthcare capacity, ii) epidemic trajectories; and iii) drug efficacy in the absence of supportive care. In each case, the outcome of interest was the number of COVID-19 deaths averted in scenarios with the therapeutic compared to scenarios without. We find the impact of drugs like dexamethasone (which are delivered to the most critically-ill in hospital and whose therapeutic benefit is expected to depend on the availability of supportive care such as oxygen and mechanical ventilation) is likely to be limited in settings where healthcare capacity is lowest or where uncontrolled epidemics result in hospitals being overwhelmed. As such, it may avert 22% of deaths in high-income countries but only 8% in low-income countries (assuming R=1.35). Therapeutics for different patient populations (those not in hospital, early in the course of infection) and types of benefit (reducing disease severity or infectiousness, preventing hospitalisation) could have much greater benefits, particularly in resource-poor settings facing large epidemics.

**Conclusions:** There is a global asymmetry in who is likely to benefit from advances in the treatment of COVID-19 to date, which have been focussed on hospitalised-patients and predicated on an assumption of adequate access to supportive care. Therapeutics that can feasibly be delivered to those earlier in the course of infection that reduce the need for healthcare or reduce infectiousness could have significant impact, and research into their efficacy and means of delivery should be a priority.

## Introduction

The COVID-19 pandemic remains a major global health threat, with 165 million cases and 3.4 million deaths confirmed worldwide as of 8th March 2021. Though characterised by extensive asymptomatic or non-life-threatening infection (1), a small proportion of infected individuals go on to develop respiratory and systemic illness requiring hospitalisation, and in critical cases, advanced respiratory support such as mechanical ventilation (2) - disease severity and risk typically increases with age. The pandemic has exerted substantial pressure on healthcare systems, with demand for oxygen, advanced respiratory support (ARS) and beds nearing or eclipsing availability in a range of settings hit hardest by the virus (3). Such pressure has been associated with increased mortality of hospitalised individuals (4). Emergence of novel SARS-CoV-2 variants with increased transmissibility (5) and/or the ability to evade immunity from previous infections (6–8) and reduce the effectiveness of vaccines (9) has underscored the potential for the virus to become endemic (10) and the need for an integrated, long-term approach to combating COVID-19.

Many clinical trials and other studies have been initiated to evaluate potential therapeutics. Dexamethasone, a corticosteroid, has been shown to reduce mortality in both severely/critically-ill (11) and moderately ill patients (12), is now recommended for use by the World Health Organisation (WHO) and used widely (13). Recent evidence also indicates the potential efficacy of therapeutic anti-coagulation (as severely ill COVID-19 patients frequently present with coagulation abnormalities (14)) in some patients (15), as well as interleukin-6 receptor antagonists, such as tocilizumab and sarilumab (16). Other candidates have included antivirals such as Remdesivir (selected for its ability to potentially suppress viral loads, which have been correlated with COVID-19 severity and mortality (17–19)), although its effect remains uncertain (20, 21). Recent months have also seen trials focussed on individuals who are not hospitalised, including those aiming to prevent progression to hospitalisation, such as for colchicine (22) and monoclonal antibodies (23, 24), with the latter also hypothesised to reduce viral loads (and so, potentially, infectiousness).

This array of potential COVID-19 therapeutics spans a range of different epidemiological impacts (from reductions in mortality through to impacts on healthcare demand and community transmission) as well as the patient populations to whom they are administered (e.g. hospitalised individuals, outpatients or those with only mild symptoms). In the context of these diverse properties, understanding the potential impacts of each, and how this is affected by other factors (epidemic context and healthcare supply) is vital for guiding procurement and research priorities. Here we use a modelling approach to understand the impact of established and potential COVID-19 therapeutics on disease burden and how this is affected by epidemic context and healthcare resources. Our results highlight how limited healthcare resources can constrain therapeutic impact, limiting the benefits of existing therapeutics, and provide insight into the types of properties of new therapeutics that could be of greatest value.

## Results

### Evaluating the Impact of Dexamethasone Under Different Assumptions of Epidemic Spread and Health System Capacity

Using an age-structured mathematical of SARS-CoV-2 transmission and COVID-19 disease, we simulated an epidemic in a setting with a profile typical of LMICs (age-structure identical to the LMIC with the median percentage of those >65 years, median hospital beds per capita and assumptions surrounding the availability of oxygen and advanced respiratory support, in-keeping with recent estimates highlighting limited availability (25, 26) - see Supplementary Information for further information) under two epidemic scenarios (R = 1.35 or 2). Our results highlight the substantial difference in the timing and intensity of healthcare demand resulting from epidemics of different sizes. Higher R epidemics (R = 2, representing a poorly mitigated epidemic) lead to a smaller fraction of moderately ill patients (requiring a general hospital bed, **Fig.2A)** and severely/critically ill patients (requiring ICU-based care, **Fig.2B)** receiving the clinical care they need, with this disparity most pronounced for ICU-based care. A lower R reduces demand for healthcare, resulting in a higher proportion of individuals receiving the required care, but still leaves a high proportion not receiving the full ICU-based care they need. Given these differences in the proportion of individuals receiving healthcare under different epidemic scenarios, we next examine the ‘Infection Fatality Ratio’ (i.e. the probability of death given infection) that persons with SARS-CoV-2 face, as a result of the joint effect of disease, healthcare capabilities and the possible usage of dexamethasone. Our results highlight the pronounced impact of healthcare constraints on the infection fatality ratio (IFR), which is significantly higher when healthcare resources (ARS, O2 and beds) are limited (**Fig. 2D**: dots). This increase in the IFR is most substantial for our high R scenario where more intense epidemics result in a higher fraction of individuals not receiving adequate care.

**Figure 1:**
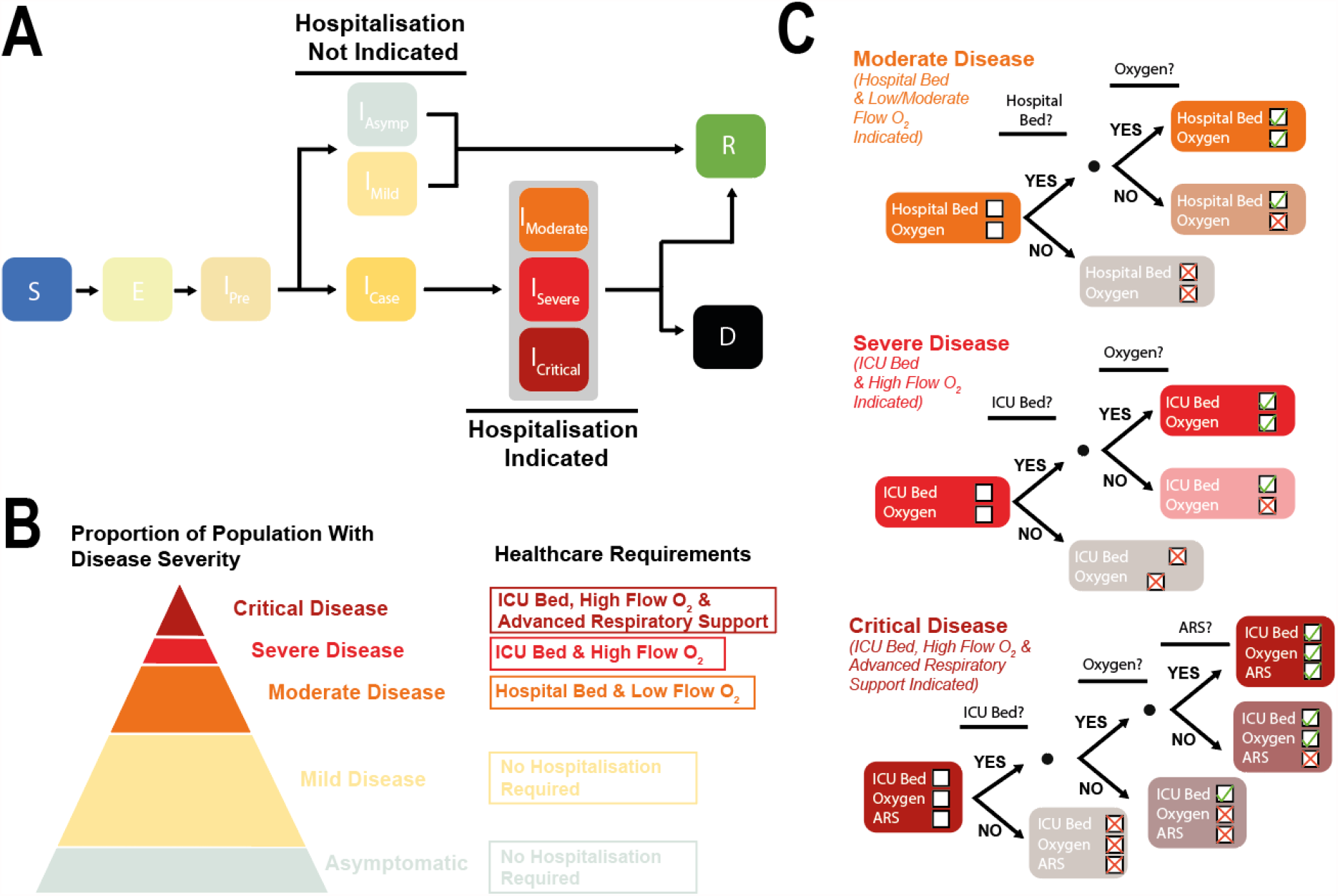
Mathematical modelling approach used to evaluate potential COVID-19 treatment impact. **(A)** Schematic representation of the natural history of SARS-CoV-2 infection and COVID-19 disease in the model. **(B)** Description of the different disease states included in the model and the associated healthcare requirements. **(C)** Decision-tree diagrams illustrating the conditional delivery of healthcare components according to disease severity and availability. There is excess mortality associated with not receiving the full set of required healthcare components.

**Figure 2:**
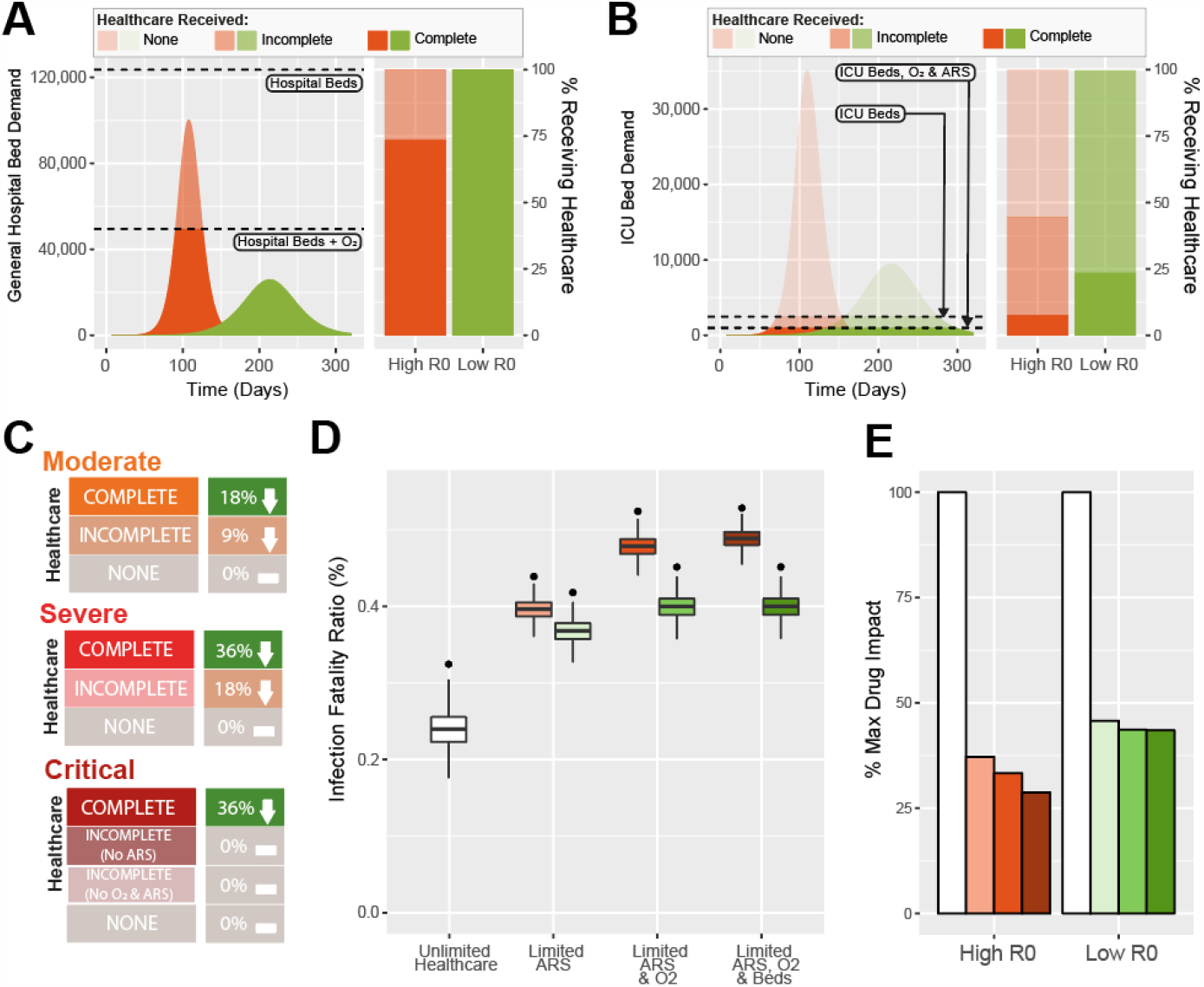
Projected impact of Dexamethasone on COVID-19 mortality under different scenarios of epidemic progression and healthcare availability. **(A)** Daily general hospital bed demand under an epidemic scenario with a high reproduction number (R = 2, orange) or a low reproduction number (R = 1.35, green). Dashed lines indicate availability of different healthcare resources, and the right hand panel describes the proportion of patients that require oxygen and a general hospital bed who receive complete (bed and oxygen), incomplete (bed only) or no healthcare (neither). **(B)** As for **(A)**, but describing demand and healthcare received for severely and critically ill patients requiring an ICU bed, oxygen and advanced respiratory support (ARS). **(C)** Schematic illustration of the impact assumed for dexamethasone on COVID-19 mortality in different patient populations (moderate, severe or critical illness), and according to the care received (complete, incomplete or none) **(D)** The impact of Dexamethasone on the COVID-19 infection fatality ratio under different assumptions for R (low, green or high, orange) and healthcare availability (unlimited, limited ARS, limited ARS and oxygen or limited ARS, oxygen and beds). In all panels, black points show the IFR without Dexamethasone, and the boxplots show the modelled IFR using the assumed Dexamethasone clinical benefit estimates described in **(C). (E)** The percentage of maximum potential Dexamethasone impact (defined as the reduction in IFR achieved by Dexamethasone under a situation of unlimited healthcare) achieved in each of the different scenarios for healthcare availability. Orange and green bars refer to high and low R scenarios respectively, with the shading indicating the extent of imposed healthcare constraints, coloured as for **(D)**.

We find that the therapeutic impact of dexamethasone (**Fig. 2D**: boxes) is strongly dependent on these same factors: there is a substantial reduction in mortality due to the drug when there are adequate healthcare resources, but a much smaller effect when these resources are unavailable. This is especially the case when a larger epidemic has overwhelmed resources **(Fig.2D)**. The reduced impact of dexamethasone in these circumstances is because fewer individuals are hospitalised and receive dexamethasone (due to shortages of beds) and fewer hospitalised individuals receive the other healthcare required (oxygen/ARS) to maximise the therapeutic benefit of dexamethasone. As a result, prevailing healthcare resources in this typical setting allow only 45% (if R is 1.35) or 28% (if R is 2.0) of the maximum potential impact of dexamethasone (defined as the reduction in IFR achieved by the drug under a scenario of no healthcare resource constraints) to be realised (**Fig. 2E**).

We distinguish two layers of uncertainty in characterizing the effect of Dexamethasone: the magnitude of the effect when supportive care is available (corresponding to the findings in trials (11, 12)), and the extent to which these effects would be found for patients without such care (for which the first of these is well understood and represented in the box plots of Fig. 2D. The second is not well understood but we constructed three alternative scenarios for the extent to which the patients without supportive care may benefit from Dexamethasone, based on clinical input described in the **Supplementary**). Overall, we find that there could be only a small extra impact (with 60% of potential drug impact realised under a low R scenario, and 52% under a high R scenario) if it was assumed that individuals for whom supportive care could not be provided still benefited to some degree from Dexamethasone **(Supp Fig.3)**.

### Evaluating the Potential Impact of Dexamethasone Globally

Next, we aimed to understand how these constraints will modulate the potential impact of dexamethasone globally. We find the limitations in healthcare capabilities that limit dexamethasone’s impact are likely to be most severe in lower and middle income countries. This outweighs the opposite effect of those populations being younger on average, which would tend to lead to fewer COVID-19 hospitalisations and deaths. Under scenarios where extensive mitigation of transmission is achieved globally (defined as leading to an R of 1.35, **Fig.3A**), we expect a median of 28%, 43%, 91% and 100% of Dexamethasone’s maximum potential impact to be achieved across LICs, LMICs, UMICs and HICs respectively **(Fig.3B)**, corresponding to averting 8%, 13%, 20% and 22% of total deaths respectively. By contrast, under scenarios where epidemics are less controlled (defined as R equal to 2, **Fig.3C**), this reduces to 18%, 26%, 43% and 71% of Dexamethasone’s maximum potential impact (and 5%, 7%, 13% and 18% of total deaths averted) **(Fig.3D)**. These results highlight that innovations in therapeutics available currently will not yield equitable impact globally. This is especially concerning given that the same countries in which existing therapeutics are predicted to have limited benefit are also those expected to see much later and slow scale-up of vaccines; where epidemic control is least sustainable; and where the emergence of novel variants (which are not explicitly accounted for in the developed framework) is currently leading to rapid resurgence of transmission (such as across Southern Africa (27) and Brazil (8, 28)).

**Figure 3:**
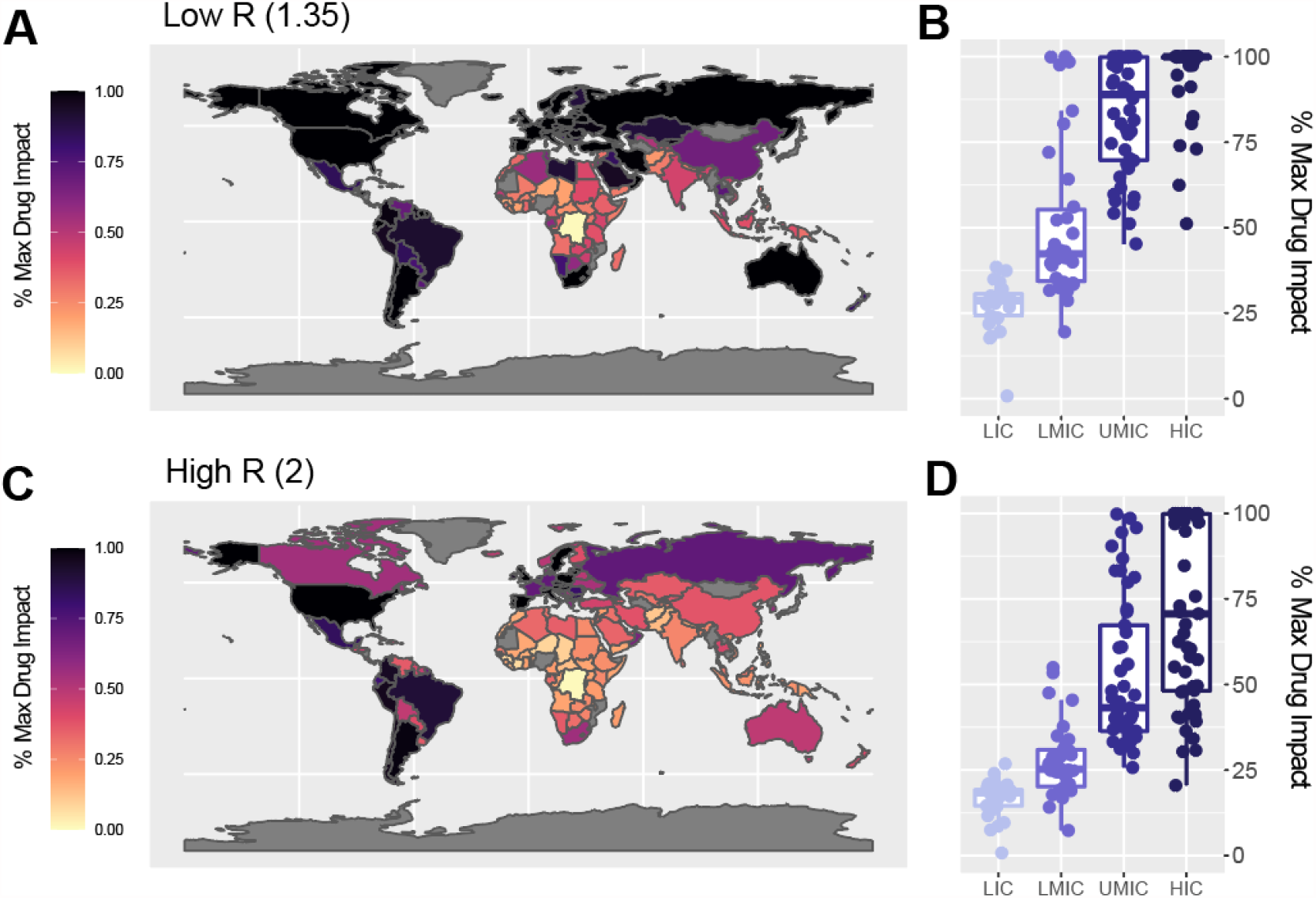
The global impact of Dexamethasone on COVID-19 mortality under different assumptions for future transmission and epidemic spread. **(A)** The percentage of maximum potential Dexamethasone impact (defined as the reduction in IFR achieved by Dexamethasone under a situation of unlimited healthcare) achieved for each country under an epidemic scenario of extensive mitigation control (R = 1.35). **(B)** The percentage of maximum Dexamethasone impact achieved in each country. Each dot is the result for a single country, coloured according to the World Bank strata that country belongs to, with the boxplot presenting summary statistics for the modelled countries in aggregate. **(C)** As for A, under an assumption of an epidemic scenario characterised by uncontrolled spread (R = 2). **(D)** As for B, under an assumption of an epidemic scenario characterised by uncontrolled spread (R = 2).

### Exploring the Potential Impact of Different Treatments and Drug Properties

We divide the spectrum of potential effects of therapeutics currently under investigation into six types (see **Table 1**), and explore their capacity to avert COVID-19 mortality under a range of scenarios varying potential effectiveness, coverage and epidemic intensity (low or high R, see **Fig.4A** for low R scenario results and **Supp Fig.4** for results for high R scenario results). The impact of therapeutics administered to hospitalised patients (types 1, 2 and 3) have a lower overall impact in reducing deaths overall, even when efficacy and coverage are high, because they suffer from the limitation that their therapeutic benefit is dependent on similar healthcare capabilities (such as the availability oxygen and ARS) described above for dexamethasone **(Fig.4A, top row)**. Therapeutics that reduce severity of disease (Type 2) or reduce the duration of hospitalisation (Type 3) do have an indirect effect in alleviating healthcare demand, but this is found to be small. The indirect effect is small in these conditions because demand for healthcare resources outstrips supply by such a wide margin, even under comparatively well-controlled epidemics (low R scenario), that a slightly faster throughput of patients does not substantially reduce the number of those that do not access healthcare at all.

**Table 1:** Potential COVID-19 therapeutic effects and their impacts. (Note: Inclusion in this list indicates that studies are underway to test for this property, and not that evidence has been found.

| Effect | Description | Target Population | Epidemiological Impact | Examples of Therapeutics Which May Have This Property* | Indicative Potential Efficacy Range | Indicative Potential Coverage Range |
| --- | --- | --- | --- | --- | --- | --- |
| Type 1 | Reduce COVID-19 mortality | Hospitalised patients (moderately, severely or critically ill) | Reduced mortality | Dexamethasone (moderately(12) & severely/critically ill patients(11, 12))<br>Remdesivir (moderately ill patients(40, 41), per the meta-analysis of the two trials)<br>Tocilizumab and Sarilumab (severely/critically ill patients(16))<br>Therapeutic anticoagulants (moderately ill patients(15)) | 20-45% relative reduction in mortality | 90-100% |
| Type 2 | Reduce COVID-19 severity (in hospitalised patients) | Hospitalised patients (moderately, severely or critically ill) | Reduced mortality and healthcare pressure | Possibly therapeutic anticoagulants (moderately ill patients(15)) | 20-45% relative reduction in hospitalised patients requiring ICU stay | 90-100% |
| Type 3 | Reduce duration of hospitalisation with COVID-19 | Hospitalised patients (moderately, severely or critically ill) | Reduced healthcare pressure | Remdesivir (moderately ill patients(40)) | 20-45% decrease in duration of hospitalisation | 90-100% |
| Type 4 | Prevent hospitalisation due to COVID-19 | Post-symptom onset. Mildly symptomatic individuals in the community | Reduced mortality and healthcare pressure | Monoclonal antibodies(23, 24)<br>Colchicine(22) | 25-75% reduction in chance of hospitalisation | 25-50% |
| Type 5a | Reduce duration of infectiousness | Post-symptom onset. Mildly symptomatic individuals in the community | Reduced mortality, healthcare pressure and transmission | Postulated for Monoclonal antibodies due to effect on viral loads(23, 24) | 25-75% reduction in duration of infectiousness | 25-50% |
| Type 5b | Reduce duration of infectiousness | Post-exposure. All individuals exposed to risk of infection, irrespective of symptoms | Reduced mortality, healthcare pressure and transmission | Postulated for Monoclonal antibodies due to effect on viral loads(23, 24) | 20-75% reduction in duration of infectiousness | 10-25% |

**Figure 4:**
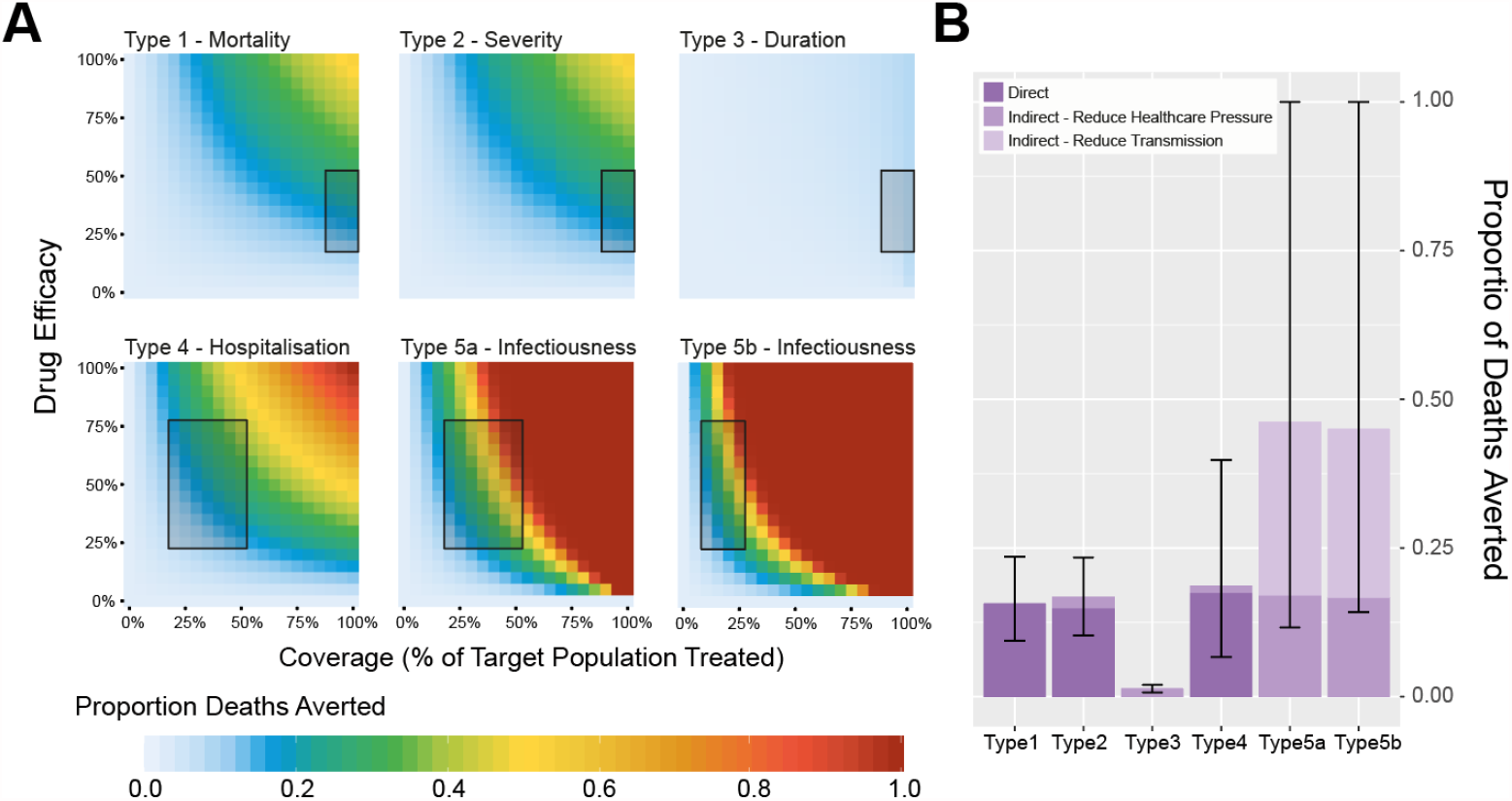
Impact of different therapeutic product effects on COVID-19 disease burden. **(A)** For an epidemic with an R of 1.35, the proportion of COVID-19 deaths averted as a function of therapeutic efficacy and therapeutic coverage, for 6 different types of potential effects (Table 1). These include reducing COVID-19 disease mortality (Type 1); preventing deterioration and worsening of disease in hospitalised patients (Type 2); reducing duration of hospitalisation (Type 3); preventing hospitalisation due to COVID-19 (Type 4) and reducing duration of infectiousness, either among symptomatics (Types 5a) or all infected-persons (Type 5b). Inset boxes indicate the range of plausible values of coverage used to generate the estimates in **(B). (B)** Disaggregation of therapeutic effect type impact by whether this is direct or indirect. Bars are coloured according to the type of impact (direct reduction in mortality, indirect reduction in mortality due to reduced pressure on healthcare or indirect reduction in mortality due to reductions in community transmission), with error bars indicating the maximum and minimum proportion of deaths averted under the range of coverage and effectiveness values considered for each effect type (indicated by the boxes in **(A)** and **Table 1**).

In comparison, therapeutics that are not administered in hospitals (and so do not suffer the same limitations) and address patients at an earlier stage of disease progression have at least a comparable, and potentially greater impact, even after allowing for the much lower coverages of such therapeutics that may be achieved **(Fig.4A, bottom row)**. Type 4 therapeutics (which reduce likelihood of severe disease and hospitalisation) have both a direct effect (in reducing mortality) and an indirect effect (through reducing healthcare demand and in doing so enabling greater access to healthcare for others) - and avert a significant fraction of COVID-19 mortality. For our low R scenario, there is an even greater effect from Type 5 therapeutics (which reduce infectiousness) through their impact on community transmission, which both reduces the overall number of people infected with SARS-CoV-2 during the epidemic and alleviates demand for healthcare resources. This would be especially the case if therapeutics are administered before onset of symptoms (Type 5b), although the coverage that could be achieved with such therapeutics would be expected to be lower than that of therapeutics administered rapidly following onset of (mild) symptoms (Type 5a). It follows that the estimates of impact are influenced by R and healthcare resources (see **Supp Fig 4** in SI) - for Type 5a and Type 5b therapeutics, relative impact is higher under the low R scenario, and lower in our high R scenario (though still comparable with the best performing hospital administered therapeutics i.e. our Type 1 profiles); for our high R scenario, Type 4 therapeutics were predicted to have the greatest benefit in the range of indicative coverages and efficacies explored **(Supp Fig 4B)**. If healthcare needs do not eclipse resources, the direct effect of hospital-delivered therapeutics is greater than otherwise, although the overall impact on mortality from Types 4, 5a and 5b remains high.

## Discussion

Understanding the contexts in which COVID-19 treatments are likely to be most effective is essential for guiding research and procurement. Here, we utilise a mathematical modelling approach to evaluate the potential impact of COVID-19 treatments under a range of different assumptions about healthcare availability and future epidemic trajectory. Overall, our results show that effect sizes for therapeutics estimated in clinical trials will not necessarily provide a guide as to their potential ‘real-world’ impact on COVID-19 disease burden. In addition to the effects measured in a randomised controlled trial setting, ‘real-world’ impact also crucially depends on prevailing healthcare constraints, the trajectory of the epidemic and the extent to which therapeutics benefits persist in the absence of supportive care. We find that, as a result, although the impact of the main therapeutic currently recommended recommended by the WHO (dexamethasone) could be considerable in well-resourced settings with an epidemic under control (averting almost a quarter of deaths), it could be far smaller in settings where resources are limited and/or there is large epidemic (averting less than 10% of deaths).

Our results also reveal that treatments with different types of effect can yield vastly different scales of population-level impact: this should guide priority setting for further investigations. In particular, the results highlight the substantial impacts that could be achieved with therapeutics delivered to persons not in hospital that either reduce the duration of infectiousness (leading to reductions in transmission) or disease severity (preventing hospitalisation and reducing healthcare strain): our results highlight that even modest levels of treatment efficacy or coverage could achieve high levels of impact. Importantly, because of the nature of their administration (delivered in the community) and the effects of these therapeutics (which do not depend on the presence of other hospital-based healthcare resources), our results suggest their potential impact would also be less affected by constraints on healthcare resources. However, this would need to be balanced by the ability of healthcare systems to deliver therapeutics in the community (including health worker capacity and distribution and storage channels) and the costs of doing so. Whilst our results have highlighted that only modest levels of coverage among patient populations with these therapeutics is required for significant impact, such levels are likely impractical for therapeutics requiring infusion such as monoclonal antibody therapies. The current cost of these therapies is also substantial and may prove prohibitive in all but the most well-resourced settings. Achieving levels of coverage required for substantial impact would instead likely prove more feasible for orally delivered, low-cost therapeutics, perhaps such as colchicine (which recent work has indicated may have the potential to reduce the need for hospitalisation, a Type 4 property (22)).

Despite this potential impact, the majority of trials to date have focussed on evaluating treatments aimed at the most critically ill and hospitalised patients. The results presented here underscore the urgent need for more trials evaluating treatments for outpatient populations and persons with mild to moderate COVID-19 disease: such as PRINCIPLE (evaluating the inhaled corticosteroid budesonide in outpatient populations) and ANTICOV (led by the Drugs for Neglected Diseases Initiative and evaluating a number of different therapeutics in 13 countries across Africa (29)). However, achieving impact with these therapeutics will be contingent on ensuring individuals receive them in a timely manner following symptom onset if the full benefits of such treatments (particularly on infectiousness and onwards transmission) are to be realised. This underscores the importance of accompanying clinical trials with operational research in order to ensure mechanisms for drug delivery to affected communities can occur in a way that the benefit is realized.

A related caveat to the results presented here is that we assume levels of healthcare seeking within the population such that all individuals with COVID-19 requiring hospitalisation will seek care. However, numerous studies have highlighted the disparities in access to healthcare that exist globally (e.g. (30, 31)), and that cost of care (if borne privately) can be a key deterrent (32). These results are consistent with an emerging body of evidence suggesting that in many low-income settings, a substantial fraction of COVID-19 deaths to date have occurred in the community (33). To the extent that not all of those in need seek care, the limitations we have found for therapeutics for hospitalised patients and the potential benefits of therapeutics for non-hospitalised patients would be even greater than our results show. The framework developed here does not incorporate waning immunity or the possibility of novel SARS-CoV-2 variants able to partially evade protective immunity elicited by prior infection (as in Brazil (8), South Africa (34) and now many other countries). These factors would serve to reduce the effect of population-level immunity and contribute to larger further waves of disease - these are also scenarios under which our findings on the influence on healthcare constraints would be magnified.

A limitation to the analyses presented here is that we do not consider differences in admission, triaging and treatment policies across different settings. In practice, there are many ways in which healthcare facilities can manage their resources so as to minimise the mismatch between supply (incoming COVID-19 patients requiring care) and demand (available healthcare resources) induced by an epidemic (e.g., alterations to triaging practices, resource sparing and alterations to criteria for admission) and these are likely to vary in response to local conditions However, we do not expect these factors to materially affect our results as any such management would not fully alleviate the large mismatch between supply and demand anticipated in these analyses (as in Figure 2). We similarly have not considered the relative costs of these different therapeutics, but our findings could be used as the basis for an assessment of their cost-effectiveness. By the same token, our results do not suggest that Dexamethasone should not be used or procured or that is not a cost-effective treatment currently.

We have found that existing global inequities in health system capacity are compounded by the action of currently available therapeutics. In fact, this could become magnified further as the rollout of COVID-19 vaccines take place, and protection from symptomatic infection and severe disease concentrates in middle- and high-income countries. Despite evidence that globally equitable vaccination strategies would be far more effective at averting mortality (35), COVID-19 vaccine doses to date have almost all been delivered to middle- and high-income countries. Initiatives aimed at equitable distribution such as the COVAX facility (part of the Access to COVID-19 Tools Accelerator initiated by the World Health Organisation) might mitigate these issues to an extent, but are unlikely to be able to facilitate high levels of population coverage in the short to medium term. Sustained epidemic control is less likely in the lowest-income population and recent resurgences observed across Latin America and parts of Southern Africa, likely attributable to emergence of novel SARS-CoV-2 variants, have highlighted the continued threat the virus poses. Taken together, this suggests a scenario in which, if further new treatments with different types of effect are not found, then not only are low and lower-middle income countries the last to access protection from the virus (in the form of vaccines), but also least able to reap the benefit of therapeutic innovations achieved to date.

## Materials and Methods

### Mathematical Model of SARS-CoV-2 Transmission

We extended a previously published model of SARS-CoV-2 transmission*(36)* to include an updated representation of COVID-19 disease, healthcare capacity and the impact of potential pharmaceutical therapeutics (See Appendix). Briefly, the model is a deterministic, age-structured elaboration of an ‘SEIR model’ **(see Fig.1A** and **Supp Fig.1)**. Upon infection, individuals move to a latent state (but are not yet infectious). Following both an incubation period and a pre-symptomatic period (in which individuals are yet to have symptoms but are infectious), individuals move to one of three disease states: asymptomatic infection, symptomatic but mild infection or infection with more serious symptoms that will eventually require hospitalisation. This process occurs in an age-dependent manner, with younger individuals more likely to be asymptomatic or have mild symptoms, and older individuals more likely to have serious symptoms. For asymptomatic or mildly symptomatic individuals, we assume there is no mortality associated with the infection, and these individuals go on to recover from the acute infection without healthcare. Those with more serious symptoms are assumed to deteriorate to the point of requiring hospitalisation, whereupon they progress to either moderate disease (requiring a general hospital bed and low/moderate flow oxygen), severe disease (requiring an ICU bed and high flow oxygen) or critical disease (requiring an ICU bed, high flow oxygen and some form of advanced respiratory support,(ARS)) **(Fig.1B)**. The model dynamically tracks healthcare resources (beds, oxygen and ARS devices) and their use in order to determine what care an individual actually receives **(Fig.1C)**. Individuals in the Moderate, Severe and Critical disease severity states then go on to either Recover or Die, with the probability of this occurring as a function of an individual’s age, disease severity and healthcare received. See the Appendix for further information on model parameterisation and a detailed description of the model formulation.

### Model Parameterisation

Natural history parameters for SARS-CoV-2 infection were taken from the literature where possible (see **Supplementary Tables 1-3**). Clinical parameters such as the duration of hospital stay were derived from a literature review of publications spanning 20 countries (See **Supplementary Table 4**), and the overall infection fatality ratio (IFR) was derived from Brazeau et al*(37)*. To derive estimates for certain parameters we convened a clinical panel of 34 medical professionals who have treated patients with COVID-19 in 11 countries (Argentina, Brazil, Colombia, Ecuador, India, Indonesia, Kenya, Thailand, United Kingdom, Venezuela and Zambia). This focused on determining i) the potential effect of dexamethasone under different assumptions of healthcare availability and ii) the overall effect of healthcare resource unavailability (either lack of advanced respiratory support, oxygen or beds) on COVID-19 mortality. See Supplementary Information for collated responses and further information about how these were integrated into model parameterisation.

### Model Simulation

To evaluate the impact of different therapeutics on COVID-19 mortality, we simulated epidemics under varying degrees of healthcare availability and epidemic trajectories. We first simulated an epidemic in a setting with a profile typical of lower-middle income countries (with a population age-structure equivalent to the lower-middle income country with the median percentage of those >65 years and median hospital beds per capita for this income strata) under two epidemic scenarios that reflected different extents of control; specifically a scenario with a high reproduction number for a poorly mitigated epidemic (R = 2), and another with a low reproduction number for a partially mitigated epidemic (R = 1.35). We also vary the availability of healthcare resources, exploring scenarios with i) unlimited healthcare, ii) where the availability of advanced respiratory support (ARS) only is limited, iii) where ARS and oxygen availability are both limited; and iv) where ARS, oxygen and hospital/ICU beds are all limited. For country-specific estimation, we fit our model to officially reported COVID-19 deaths data collated by Worldometer*(38)* and the Johns Hopkins Coronavirus Resource Centre*(39)* and project the epidemic forwards under different assumptions of future epidemic control (specifically either a high or low reproduction number as described above). Model fitting was carried out within a Bayesian framework, using an adaptive Markov-Chain Monte Carlo (MCMC) scheme and statistical inference based on the results from 20,000 iterations. To evaluate the potential impact of different therapeutics, we consider 6 different types of therapeutic effects, each of which is hypothesized to be a mode of action of at least one proposed therapeutic (see Table 1). For each, we proposed indicative ranges for efficacy and the achievable coverage of the relevant patient population as a mean to benchmark comparisons drawn between their respective effects.

## Data Availability

All data and outputs relevant to the analysis are available at https://github.com/mrc-ide/apothecary.

## Author Contributions

CW & TBH conceived the study. CW, OJW and PGTW undertook the modelling and data analysis, with input from TBH, ACG, AH, HT, PW and AQ. All other authors contributed to forming modelling assumptions and interpreting results in a clinical panel, or assisted in the setting up of the clinical panel. CW & TBH produced the first draft of the manuscript. All authors contributed to the final draft.

## Data and Materials Availability

All data, code and materials used in this analysis are available at https://github.com/mrc-ide/apothecary.

## Supplementary Materials

- **Supplementary Materials and Methods**
- **Figure S1:** Age-structured model of SARS-CoV-2 transmission explicitly incorporating disease severity, healthcare capacity and passage through different healthcare levels.
- **Figure S2:** Decision trees dictating healthcare individuals receive, for each of the disease severity categories modelled within the framework.
- **Figure S3:** The effect of varying assumptions surrounding dexamethasone’s impact on COVID-19 mortality in the absence of other healthcare.
- **Figure S4:** Impact of different therapeutic product effects on COVID-19 disease burden, for a high R scenario.
- **Supplementary Results:** Model Fitting Results for All Countries

