## Supplementary Information for "Understanding the Potential Impact of Different Drug Properties On SARS-CoV-2 Transmission and Disease Burden: A Modelling Analysis"

[1]MRC Centre for Global Infectious Disease Analysis, Imperial College London, London, United Kingdom
[2]Clínica Universitaria Colombia, Clínica Colsanitas. Facultad de Medicina, Universidad Nacional de Colombia, Bogotá, Colombia
[3]Division of Infectious Diseases and Tropical Medicine, Department of Medicine, Faculty of Medicine Siriraj Hospital, Mahidol University, Bangkok, Thailand
[4]Faculty of Medicine Siriraj Hospital, Mahidol University, Bangkok, Thailand
[5]Hospital Universitario San Ignacio -Pontificia Universidad Javeriana, Bogotá, Colombia
[6]Adult Infectious Diseases Centre, University Teaching Hospital, Lusaka, Zambia
[7]Department of Internal Medicine, University of Zambia School of Medicine, Lusaka, Zambia
[8]Bamrasnaradura Infectious Diseases Institute, Department of Diseases Control, Ministry of Public Health, Nonthaburi, Thailand
[9]Department of Infectious Diseases, Imperial College London, London, United Kingdom
[10]NIHR Biomedical Research Centre, Imperial College NHS Trust, London, United Kingdom
[11]Oswaldo Cruz Foudantion, Mato Grosso do Sul, Campo Grande, Brazil
[12]School of Medicine, Federal University of Mato Grosso do Sul, Campo Grande, Brazil
[13]Yale School of Public Health, New Haven, USA
[14]Faculty of Medicine, Pontificia Universidad Javeriana, Bogotá, Colombia
[15]Eijkman-Oxford Clinical Research Unit, Jakarta, Indonesia
[16]Faculdade de Medicina de São José do Rio Preto (FAMERP), São José do Rio Preto, Brazil
[17]Instituto de Zoologia y Ecologia Tropical, Facultad de Ciencias, Universidad Central de Venezuela, Caracas, Venezuela
[18]Departamento de Molestias Infecciosas e Parasitarias and Instituto de Medicina Tropical da Faculdade de Medicina da Universidade de São Paulo, São Paulo, Brazil
[19]Department of Zoology, University of Oxford, Oxford, UK.
[20]Hospital das Clínicas da Faculdade de Medicina da Universidade de São Paulo, São Paulo, Brazil
[21]Biomedical Research and Therapeutic Vaccines Institute, Ciudad Bolívar, Venezuela.
[22]MRC Clinical Trials Unit at University College London, London, United Kingdom
[23]Division of Anaesthetics, Pain Medicine and Intensive Care, Imperial College London, London, United Kingdom
[24]Centre for Tropical Medicine and Global Health, Nuffield Dept of Medicine, University of Oxford, Oxford, UK
[25]Fatmawati General Hospital, Faculty of Medicine University of Indonesia
[26]Samutprakan Hospital, Bangkok, Thailand
[27]Kenyan Ministry of Health, Nairobi, Kenya
[28]Instituto Venezolano de Investigaciones Científica, Caracas, Venezuela
[29]Department of Infectious Diseases, Faculdade de Medicina, Universidade de São Paulo, São Paulo, Brazil
[30]Hospital Bernardo Houssay, Buenos Aires, Argentina
[31]Department of Epidemiology and Biostatistics, Imperial College London, London, United Kingdom
[32]Department of Clinical Tropical Medicine, Faculty of Tropical Medicine, Mahidol University, Bangkok, Thailand
[33]Kenyan Ministry of Health, Kiambu County, Kenya
[34]OneHealth Global Research Group, Universidad de las Américas, Quito, Ecuador
[35]School of Medical Sciences; University of Campinas, Campinas, Brazil
[36]Institute for Global Health, University College London, London, United Kingdom
[37]Critical Care Division, Department of Medicine, Faculty of Medicine Siriraj Hospital, Mahidol University, Bangkok, Thailand
[38]School of Public Health, Imperial College London, London, United Kingdom
[39]Instituto de Medicina Tropical da Faculdade de Medicina da Universidade de São Paulo, São Paulo, Brazil
[40]Section of Adult Infectious Disease, Department of Infectious Disease, Faculty of Medicine, Imperial College London, London, UK
[41]Department of Anesthesiology and Intensive Theraphy, Faculty of Medicine, Public Health and Nursing Universitas Gadjah Mada. Public Hospital Dr. Sardjito, Yogyakarta
[42]Atherosclerosis and Vascular Biology Laboratory, State University of Campinas, Campinas, Brazil
[43]Bangkok Christian Hospital, Bangkok, Thailand
[44]Hinduja Hospital and Research Centre, Mumbai, India
[45]Division of Infectious Diseases. School of Medicine. Pontificia Universidad Javeriana, Hospital Universitario San Ignacio. Bogotá, Colombia.

### Contents


#### Supplementary Materials & Methods

##### Data Sources

- Contact Matrices
- Intervention and Mobility Data
- Demographic Data
- Healthcare Capacity Data
- Mortality Data
- Clinical Panel Input

##### Mathematical Model

- Model Description
- Model Equations
- Model Fitting
- Parameter Description and Values

#### Supplementary Figures

- **S1:** Transmission Model Schematic
- **S2:** Decision Trees for Dynamically Allocating Healthcare Resources
- **S3:** Effect of Different Dexamethasone Assumptions On Impact
- **S4:** High R Results for Different Therapeutic Types

#### References

### Materials and Methods

#### Data Sources

##### Demographic

Population sizes and the proportion of individual across different age groups by country were collated from the 2020 World Population Prospects database: https://population.un.org/wpp/, the 27th round of the official United Nations population estimates that are prepared annually by the Population Division of the United Nations Secretariat.

##### Contact Matrices

Patterns of contact between different age-groups and across different countries were collated from a variety of sources. These include an update to a recent systematic review of social contact surveys to include lower and lower-middle income countries, the results of which are presented in Walker et al. Full details of the review process are available in that reference. In addition to these surveys, we also included all those downloadable via the socialmixR package (https://github.com/sbfnk/socialmixr). This full set of contact matrices from a variety of different sources are available for download as part of an R package for an SEIR model of SARS-CoV-2 transmission (https://github.com/mrc-ide/squire, the precursor to the model presented here).

##### Healthcare Capacity – Hospital Beds

Data on the number of hospital beds per 1,000 population were available from the World Bank (https://data.worldbank.org/indicator?tab=all) for 201 countries (66 High Income, 58 Upper Middle Income, 47 Lower Middle Income and 30 Low Income). Countries varied widely in how recent their most recent entry was (many were earlier than 2015). Following the approach described in Walker et al[1], we use a boosted regression tree (BRT)-based approach to generate estimates of hospital beds per 1,000 population for 2020, carried out using R statistical software and the dismo R package[2], with tree complexity of 12, bag fraction of 0.65, and a learning rate of 0.001.

##### Healthcare Capacity – ICU Beds

As with Walker et al, intensive care unit capacity data were derived from 3 resources. Data was extracted from both a previously published systematic review of ICU capacity in low-income countries[3], as well as a review of ICU capacity across Asia[4]. These data were supplemented with the results of a further systematic review, the full details of which are available in Walker et al. Together, these resources provide a total of 57 data points describing the number of ICU beds per 100 hospital beds across countries belonging to the World Bank's 4 income strata (LIC, LMIC, UMIC and HIC) – in the absence of country specific information in most cases, all country-specific modelling analyses presented here use estimates of median ICU beds based on the World Bank income strata each country belongs to.

##### Healthcare Capacity – Oxygen, Mechanical Ventilator and Other Advanced Respiratory Devices Availability

Unfortunately, comprehensive, standardised estimates of these healthcare resources globally do not exist, and we therefore make the following income-strata specific assumption about oxygen/ARS availability:

- Low Income Countries: 20% of hospital/ICU beds have access to adequate oxygen (low-flow for hospital beds, high flow for ICU beds) and ARS (for ICU beds)
- Lower-Middle Income Countries: 40% of hospital/ICU beds have access to adequate oxygen/ARS.
- Upper-Middle Income Countries: 60% of hospital/ICU beds have access to adequate oxygen/ARS.
- High-Income Countries: 100% of hospital/ICU beds have access to adequate oxygen/ARS.

This assumption is in-keeping with recent estimates from the World Health Organisation about the likely unmet need for oxygen.

#### Mortality Data

Reported daily mortality incidence data for each country was taken from Worldometer[5] and the Johns Hopkins Coronavirus Resource Centre[6]. In a small number of instances, retrospective reallocation of a recorded day's deaths to earlier days lead to incidence on that day being reported as negative. In this case, we set that day's incidence to 0.

#### Individual and Governmental-Level Responses/Interventions

In order to gather information on the responses to the epidemic occurring in each country we gathered information on individual level mobility the from publicly available Google Mobility data: https://www.google.com/covid19/mobility/, which provides data on movement for each country in a number of different categories (Grocery & Pharmacy, Parks, Transit Stations, Retail & Recreation, Residential, and Workplaces). We assume that reductions in mobility across the different settings will reduce the number of potential infective contacts occurring, but that associated increases in Residential mobility does not lead to an increase in household contacts. We therefore assume that changes in transmission over time in a given setting can be summarised by averaging the mobility trends over all categories except for Residential and Parks (where for the latter we assume the contacts made outdoors contribute negligibly to transmission).

Google Mobility data are unavailable for a number of countries, and in order to infer likely patterns of mobility across countries lacking this data, we trained a Boosted-Regression Tree model on data from the publicly available ACAPs Humanitarian Database: https://data.humdata.org/dataset/acaps-covid19-government-measures-dataset) that describes government interventions employed to date. This model is trained on the timing, frequency and types of non-pharmaceutical interventions (such as travel restrictions, school closures etc) employed by each country, as well as additional sources of government interventions not listed in ACAPs sourced from the WHO PHSM database. The World Bank income status of the country was also used as a covariate. In a similar manner to the BRT model estimating hospital beds, model fitting was carried out using R and the dismo R package, with parameters as follows: tree complexity of 8, bag fraction of 0.5, and a learning rate of 0.05. 5-fold cross-validation was implemented to assess overfitting, and error associated with the test and training datasets found to be similar. The inferred mobility is then normalised such that pre-epidemic mobility is equal to 100%.

#### Clinical Opinion Panel

To derive estimates for key parameters determining i) the effects of dexamethasone under different assumptions of healthcare availability and quality and ii) the overall effect of healthcare unavailability on COVID-19 mortality, we convened a clinical panel of 34 medical professionals spanning 11 countries (Argentina, Brazil, Colombia, Ecuador, India, Indonesia, Kenya, Thailand, United Kingdom, Venezuela and Zambia). Each of these clinicians were asked to fill in a survey containing the following questions to guide parameterisation of the model, as well as assess the extent of consensus on the impact of different therapeutics under different scenarios of healthcare availability.

1. For patients with moderate disease (which we define as those requiring low/moderate flow oxygen), what level of mortality (increased risk, RR) would you expect if oxygen was NOT available, compared to if it was? Currently we have 60% of moderate disease patients dying if they do not receive oxygen.
2. For patients with severe disease (requiring high flow oxygen), what level of mortality (increased risk, RR) would you expect if oxygen was NOT available, compared to if it was? Currently we have 95% of severe disease patients dying if they do not receive oxygen.
3. For patients with disease serious enough to require hospitalisation, would you expect a reduction in mortality associated with being hospitalised (i.e. receiving general clinical care and a bed) even if oxygen and advanced respiratory support is not available?
    a. If yes, what reduction in mortality would you expect compared to those not receiving a bed and general clinical care?
4. Would you expect any therapeutic benefit of Dexamethasone in patients requiring but not receiving therapeutic oxygen?
5. Would you expect any therapeutic benefit of Dexamethasone in patients requiring but not receiving advanced respiratory support (such as mechanical ventilation)?

Opinion was collated and used to parameterise the model's assumptions surrounding i) drug impact under different scenarios of healthcare availability and ii) the impact of healthcare availability on baseline COVID-19 mortality in the absence of pharmaceutical therapeutics. A summary of this collated opinion can be found in **Supplementary Tables 1 and 2**.

### Mathematical Model

#### Overview

We extended an existing, previously published model of SARS-CoV-2 transmission[1] (Walker et al, 2020) to include more granular representation of COVID-19 disease, healthcare capacity and the impact of potential pharmaceutical therapeutics. The model is available as an open-source, freely available R package and can be downloaded from: https://github.com/mrc-ide/apothecary. Briefly, the model is a deterministic, age-structured elaboration of an SEIR model, parameterised to match best estimates of key parameters determining the dynamics of SARS-CoV-2 spread and the natural history of COVID-19 disease. It includes:

- Patterns of contact between age-groups derived from empirical contact surveys and applied in a setting (country) specific manner (where data is available).
- Explicit representation of age-dependent disease severity, as well as the associated healthcare requirements for each disease severity strata.
- Explicit representation of healthcare capacity, specifically, the availability of:
    - Hospital and ICU beds.
    - Oxygen (both low-flow and high flow).
    - Advanced respiratory support (including but not restricted to mechanical ventilators).
- Excess mortality associated with not receiving adequate clinical care (parameterised using the results of the clinical input panel described above).
- Explicit representation of pharmaceutical therapeutics such as dexamethasone, parameterised using estimates from published literature.
- Integration of Google Mobility and ACAPs humanitarian database data describing the impact of individual-level and governmental responses to COVID-19 to date. These data are incorporated alongside COVID-19 mortality data into a Bayesian model fitting framework in order to calibrate the model to the (different) epidemics experienced by each country to date.

#### Model Description

Individuals begin in a Susceptible (S) state and move to a latently infected (E) state upon infection. They then progress to one of three different disease states: $I_{Asymp}$ (asymptomatic infection), $I_{Mild}$ (symptomatic but mild infection) or $I_{Case}$ (more serious symptoms that will eventually require hospitalisation). This process occurs in an age-dependent manner, with younger individuals more likely to be asymptomatic or have mild symptoms, and older individuals more likely to have serious symptoms. For asymptomatic or mildly symptomatic individuals, we assume there is no mortality associated with the infection, and these individuals go on to recover from the acute infection. Those with more serious symptoms are modelled as deteriorating to the point of requiring hospitalisation and clinical care. Upon deterioration, individuals progress to either $I_{Mod}$ (moderate disease requiring a general hospital bed and low/moderate flow oxygen), $I_{Sev}$ (severe disease requiring an ICU bed and high flow oxygen) and $I_{Crit}$ (critical disease requiring an ICU bed, high flow oxygen and some form of advanced respiratory support such as invasive mechanical ventilation or non-invasive ventilation like biPAP or cPAP). Again, this occurs in an age-dependent manner, such that older individuals have a higher chance of developing Severe and Critical disease. For further details see **Supplementary Figure 1**.

Within the model, we explicitly track the availability of several healthcare components: hospital beds, ICU beds, oxygen and advanced respiratory support (ARS) devices. Whilst it is disease severity (Moderate, Severe or Critical) that dictates what clinical care an individual requires, it is healthcare availability that dictates what clinical care an individual actually receives. Upon progression to Moderate, Severe or Critical disease, we dynamically evaluate the occupancy of beds and the availability of oxygen and ARS devices; in turn, we assign these healthcare components to individuals in each disease severity class where available. Depending on their disease severity and availability of healthcare components, individuals are cascaded through a series of decision trees that dictate what extent of care is received **(Supplementary Figure 2)**. The result is that a subset of individuals in a given disease severity class (e.g. Moderate) will receive the full suite of required care (i.e. a general hospital bed and low/moderate flow oxygen); a subset will receive incomplete care (i.e. a general hospital bed but no oxygen); and a subset will receive no care whatsoever (i.e. no hospital bed and no oxygen). The exact proportion of individuals receiving these different extents of clinical care will dynamically vary across the course of the epidemic depending on the extent of current healthcare utilisation and availability. For all disease severity classes, we model conditional delivery of healthcare components such that, for example, receiving mechanical ventilation is dependent on mechanical ventilators being available, but also dependent on the availability of oxygen and an ICU bed. Individuals in the Moderate, Severe and Critical disease severity states then go on to either:

- Recover (R). Note that Moderate cases go straight to recovery, whilst individuals in the Severe and Critical disease states spend time in a step-down (general hospital) bed following departure from the ICU prior to recovery.
- Die ($D_{Comm}$ or $D_{Hosp}$), with the location of death determined by whether an individual was hospitalised (received a general hospital or ICU bed, $D_{Hosp}$) or not ($D_{Comm}$, for those dying in the community). The proportion of individuals dying occurs in an age, disease severity and clinical-care-dependent manner i.e. we explicitly include excess mortality associated with receiving inadequate clinical care. Over the timescales considered here, we assume immunity to be robust and sterilising.

#### Model Fitting

Using the methodological framework developed in the COVID-19 LMIC report[7], we fit the model to COVID-19 deaths data from 165 countries around the world. These time-series of daily COVID-19 deaths for each country are then integrated with information on individual-level mobility data from Google Community Mobility Reports to capture temporal variation in the stringency of control measures that result in alterations to the dynamics and effective reproduction number of the virus. During model fitting allows we allow the following parameters to vary:

1. The start date of the epidemic, $t_0$.
2. The initial $R_0$ in the absence of mobility changes, control measures or any non-pharmaceutical intervention.
3. The initial effect size of individual-level mobility on transmission, $M_\alpha$.
4. The effect size of mobility on transmission after mobility increases from its minimum, $M_\omega$ – this is incorporated in order to account for changes over time in the relationship linking mobility to the reproduction number (Ainslie et al).

a. Specifically, $M_\omega$ scales the impact of $M_\alpha$ after the minimum, such that when $M_\omega$ is equal to 1, increases in mobility after the minimum will not increase $R_t$, and when $M_\omega$ is equal to 0, there is no decoupling between mobility and transmission, such that $R_t$ will increase with increasing mobility at the same rate as it decreased with decreasing mobility prior to the minimum.

5. A number of pseudo-random walk parameters, $\rho_i$, which are introduced starting one week after the minimum in mobility, referred to as $t_m$, which serve to capture changes in transmission that are independent to mobility to reflect changes in human behaviour over time. The equation for the time-varying reproduction number is given by:

$$Rt = R0\ .f(-M\alpha.(1-M(t))\ -M\omega.M\alpha(M(t) - M(tm)) - \rho1 - \rho2\ ...\rho n)$$

Where $f(x) = = 2\ exp(x)/(1 + exp(x))$ is twice the inverse logit function, and is a formulation that has previously been used to link changes in population-level mobility to transmissibility (as measured through the reproduction number) of the virus[8]. $M(t)$ is the inferred mobility throughout the epidemic, in which 1 represents 100% mobility (i.e. no change) and 0 represents 0% mobility. To model the changing mobility independent behaviour over time, each $\rho$ parameter is set equal to 0 for each day prior to its start date. E.g., $\rho_1$is the first mobility independent change in transmission, which starts 7 days after $t_m$ and will be equal to 0 when $t < t_m + 7$. Estimated values for each of the rhos is then maintained for all future time points – for example, $\rho_1$ is then maintained for all future time points after $t < t_m + 7$. $\rho_2$ is the second mobility independent change in transmission and starts 21 days after $t_m$, i.e. 2 weeks after $\rho_1$. The overall likelihood for the model is then as follows:

$$Dt = NB(\mu.\nu,\sigma)$$

Where $Dt$ is the number of COVID-19 deaths recorded in a given country of day t, $NB$ is the Negative Binomial distribution with standard deviation, $\sigma$ and mean $\mu$ is the model predicted number of deaths occurring on day t. $\sigma$ can be expressed as $\sqrt{}\mu{+}\mu 2/r$ , where $r$ is the dispersion parameter and assumed to be equal to 2 to account for overdispersion and known inconsistencies in day-to-day reporting of deaths during the pandemic (e.g. the pronounced “weekend” effect whereby fewer deaths are recorded over weekend periods).

Model fitting and parameter inference was carried out within a Bayesian framework, using an adaptive Metropolis-Hastings Markov Chain Monte Carlo (MCMC) sampled scheme, with adaptive tuning of the proposal implemented sampling using the Johnstone-Chang optimisation algorithm[9]. For each country the model was fit to, the algorithm was run for 50,000 iterations, the first 25,000 of which was discarded as burn-in. All parameter inference results reported here are based on these 25,000 iterations. Prior distributions for our fitted parameters are given in **Supplementary Table 6** and the full fitting results for each country are presented in the **Supplementary Appendix.**

Model Parameter Description and Values

**Supplementary Table 1:** Results of Collated Clinical Opinion

| | 1. Mortality RR - Moderate Disease, No Oxygen | 2. Mortality RR - Severe Disease, No Oxygen | 3. Effect of Hospitalisation Alone on Mortality? Y/N | 3a. If Yes, RR of Bed on COVID-19 Mortality | 4. Dexamethasone Benefit in Patients Requiring But Not Receiving Oxygen? | 5. Dexamethasone Benefit in Patients Requiring But Not Receiving ARS? |
|---|---|---|---|---|---|---|
| R1 | 1.75 | 3.5 | Yes | 0.9 | Yes | No |
| R2 | 3 | 2.5 | Yes | 0.55 | No | No |
| R3 | 2.5 | 5.5 | Yes | 0.65 | Yes | No |
| R4 | 7.5 | 1.53 | No | NA | No | No |
| R5 | NA | NA | No | 1 | Yes | No |
| R6 | 1.6 | 3 | Yes | 0.9 | Yes | Yes |
| R7 | 4 | 6 | Yes | 1 | No | No |
| R8 | 3 | 6 | Yes | 0.95 | Yes | No |
| R9 | 3 | 7 | Yes | 0.7 | Yes | Yes |
| R10 | 1.5 | 2 | No | NA | Yes | Yes |
| R11 | NA | NA | No | 1 | Yes | Yes |
| R12 | 2 | 2.5 | No | 1 | Yes | Yes |
| R13 | NA | NA | Yes | 0.85 | Yes | No |
| R14 | 4 | 7.5 | Yes | 0.6 | No | No |
| R15 | 2 | NA | Yes | 0.8 | Yes | No |
| R16 | 2 | 5 | No | 1 | Yes | No |
| R17 | 2.5 | 7.5 | Yes | 0.85 | Yes | Yes |
| R18 | NA | NA | Yes | 0.85 | Yes | Yes |
| R19 | NA | 3.5 | yes | 0.75 | Yes | No |
| R20 | 3 | 3.5 | Yes | 0.7 | Yes | No |
| R21 | NA | NA | Yes | 0.8 | Yes | No |
| R22 | 15 | 20 | Yes | 0.8 | Yes | No |
| R23 | 1.75 | 2.5 | No | 1 | No | Yes |
| R24 | 1.5 | 1.5 | Yes | 0.8 | Yes | Yes |

| | | | | | | |
|---|---|---|---|---|---|---|
| **R25** | 1.5 | NA | No | 1 | Yes | No |
| **R26** | NA | NA | Yes | 1 | Yes | Yes |
| **R27** | NA | NA | NA | NA | Yes | Yes |
| **R28** | NA | 1.65 | No | NA | Yes | No |
| **R29** | NA | NA | No | 1 | Yes | No |

Note that there are fewer than 34 separate clinical opinions as a small number of clinicians filled in the questionnaire as part of a group. NA indicates non-response.

**Supplementary Table 2:** Model Parameters Derived from Clinical Opinion

| Parameter | Value | Description |
|---|---|---|
| Reduction in COVID-19 mortality from receiving a hospital bed (and associated supportive care but not including oxygen/ARS) | RR 0.85 | Derived from Clinical Opinion. |
| Increase to mortality in moderate disease if receive hospital bed but no oxygen | RR 1.91 | RR 2.25 for increased mortality associated with inadequate treatment, RR 0.85 for receiving hospital bed over no treatment at all. Derived from Clinical Opinion. |
| Increase to mortality in moderate disease if receive no hospital bed and no oxygen | RR 2.25 | RR 2.25 for increased mortality associated with inadequate treatment in moderate patients. Derived from Clinical Opinion. |
| Increase to mortality in severe disease if receive ICU bed but no oxygen | RR 2.98 | RR 3.5 for increased mortality associated with inadequate treatment, RR 0.85 for receiving hospital bed over no treatment at all. Derived from Clinical Opinion. |
| Increase to mortality in severe disease if no ICU bed and no oxygen | RR 3.5 | RR 3.5 for increased mortality associated with inadequate treatment in severe patients. Derived from Clinical Opinion. |

**Note:** For those requiring mechanical ventilation or other forms of advanced respiratory support (i.e. Critically Ill patients), but who do not receive this critically needed care, we assume either i) 95% mortality or ii) the mortality for severe disease in untreated patients, whichever is higher.

**Table S3:** Model Parameters Describing SARS-CoV-2 Transmission and COVID-19 Disease Duration

| Parameter | Value | Source/Notes |
|---|---|---|
| Basic Reproduction Number | - | Inferred from COVID-19 mortality data in a country specific manner. |
| Mean latent period | 4.6 days | Walker et al[1] |
| Mean duration of presymptomatic infection | 1.1 days | Calculated to give ~40% of transmission as presymptomatic[10] |
| Mean duration of asymptomatic infectiousness | 3.2 days | Combined duration of presymptomatic infection and mild infection (duration of asymptomatic infection assumed to be the same as mild infection) |
| Mean duration of mild infectiousness | 2.1 days | Calculated to give ~60% of transmission as post-symptoms[10] |
| Mean duration of serious infection prior to requiring hospitalisation | 4.5 days | Walker et al[1] |
| Mean duration of hospitalisation for moderate cases who die (and receive hospital bed and oxygen) | 8 days | Detailed sources for the UK[11], Brazil[12] and South Africa[13] and a rapid review of data from 15 additional countries. Described further below. |
| Mean duration of hospitalisation for moderate cases who survive (and receive hospital bed and oxygen) | 7.25 days | Detailed sources for the UK[11], Brazil[12] and South Africa[13] and a rapid review of data from 15 additional countries. Described further below. |
| Mean duration of stay in ICU for severe cases who die (and receive ICU bed and oxygen) | 9.5 days | Detailed sources for the UK[11], Brazil[12] and South Africa[13] and a rapid review of data from 15 additional countries. Described further below. |
| Mean duration of stay in ICU for severe cases who survive (and receive ICU bed and oxygen) | 6.5 days | Detailed sources for the UK[11], Brazil[12] and South Africa[13] and a rapid review of data from 15 additional countries. Described further below. |
| Mean duration of stay in ICU for critical cases who die (and receive ICU bed, oxygen and advanced respiratory support) | 11.75 days | Detailed sources for the UK[11], Brazil[12] and South Africa[13] and a rapid review of data from 15 additional countries. Described further below. |
| Mean duration of in ICU for critical cases who survive (and receive ICU bed, oxygen and advanced respiratory support) | 13.5 days | Detailed sources for the UK[11], Brazil[12] and South Africa[13] and a rapid review of data from 15 additional countries. Described further below. |
| Mean duration in recovery bed following ICU stay | 4 days | Detailed sources for the UK[11], Brazil[12] and South Africa[13] and a rapid review of data from 15 additional countries. Described further below. |

**Note:** Information on duration of hospitalisation was primarily taken from 3 sources containing detailed information on the duration of hospitalisation due to COVID-19 disease, disaggregated by disease-severity (moderate, severe or critical) and whether the individual went on to survive or die. These were derived from the UK[11], Brazil[12] and South Africa[13]. We subsequently supplemented these with the results of a rapid, non-exhaustive literature review that identified a total of 21 references from 15 additional countries (Bangladesh[14,15], China[16], France[17], Germany[18], Ghana[19], India[20], Kuwait[21], Mexico[22,23], Nepal[24], Pakistan[25], Philippines[26,27], South Africa[28], Thailand[29], Uganda[30], United Kingdom[31], USA[32] and Vietnam[33]). Estimates in the above Table represent the average of the results reported in the initial UK, Brazil and South Africa references, as well as the overall average from these other, less detailed and smaller studies.

**Table S4:** Age-Specific Model Parameters Describing COVID-19 Disease Severity

| **Age-Group (years)** | **Proportion of Infections that are Asymptomatic**[34] | **Proportion of Infections that are Hospitalised**[17] | **Proportion of Hospitalised Cases Requiring ICU Care**[17] | **Proportion of ICU Cases Requiring Advanced Respiratory Support**[11] |
|---|---|---|---|---|
| 0 to 4 | 0.30 | 0.0008 | 0.1813 | 0.7842 |
| 5 to 9 | 0.30 | 0.0012 | 0.1813 | 0.7482 |
| 10 to 14 | 0.20 | 0.0017 | 0.1813 | 0.7143 |
| 15 to 19 | 0.20 | 0.0023 | 0.1374 | 0.6884 |
| 20 to 24 | 0.20 | 0.0033 | 0.1219 | 0.6805 |
| 25 to 29 | 0.20 | 0.0046 | 0.1227 | 0.6917 |
| 30 to 34 | 0.20 | 0.0065 | 0.1360 | 0.7156 |
| 35 to 39 | 0.20 | 0.0091 | 0.1609 | 0.7451 |
| 40 to 44 | 0.20 | 0.0129 | 0.1969 | 0.7735 |
| 45 to 49 | 0.20 | 0.0180 | 0.2420 | 0.7959 |
| 50 to 54 | 0.20 | 0.0254 | 0.2893 | 0.8104 |
| 55 to 59 | 0.20 | 0.0358 | 0.3265 | 0.8166 |
| 60 to 64 | 0.20 | 0.0503 | 0.3372 | 0.8150 |
| 65 to 69 | 0.20 | 0.0708 | 0.3090 | 0.8055 |
| 70 to 74 | 0.20 | 0.0995 | 0.2437 | 0.7891 |
| 75 to 79 | 0.20 | 0.1400 | 0.1604 | 0.7669 |
| 80+ | 0.20 | 0.2335 | 0.0570 | 0.7414 |

**Table S5:** Model Parameters Describing Age and COVID-19 Disease Severity-Specific Baseline Mortality (Assuming Unlimited Healthcare and All Individuals Receive Appropriate Care)

| Age-Group (years) | Proportion of All Deaths that Occur in the ICU[11] | Proportion ICU Deaths in ARS Patients[11] | Proportion of Moderate Disease Cases Dying[35] | Proportion of Severe Disease Cases Dying[35] | Proportion of Critical Disease Cases Dying[35] |
|---|---|---|---|---|---|
| 0 to 4 | 0.2083 | 0.8654 | 0.04969 | 0.0368 | 0.0651 |
| 5 to 9 | 0.5000 | 0.8600 | 0.03494 | 0.0877 | 0.1813 |
| 10 to 14 | 0.8000 | 0.8600 | 0.01556 | 0.1378 | 0.3384 |
| 15 to 19 | 0.9500 | 0.8700 | 0.00411 | 0.2047 | 0.6197 |
| 20 to 24 | 0.9363 | 0.8722 | 0.00573 | 0.2426 | 0.7777 |
| 25 to 29 | 0.9015 | 0.8819 | 0.00987 | 0.2474 | 0.8239 |
| 30 to 34 | 0.8713 | 0.8935 | 0.01459 | 0.2350 | 0.7836 |
| 35 to 39 | 0.8502 | 0.9023 | 0.01947 | 0.2209 | 0.6983 |
| 40 to 44 | 0.8379 | 0.9055 | 0.02452 | 0.2155 | 0.6052 |
| 45 to 49 | 0.8254 | 0.9025 | 0.03116 | 0.2206 | 0.5234 |
| 50 to 54 | 0.8018 | 0.8947 | 0.04202 | 0.2319 | 0.4611 |
| 55 to 59 | 0.7589 | 0.8840 | 0.06008 | 0.2467 | 0.4223 |
| 60 to 64 | 0.6914 | 0.8714 | 0.08701 | 0.2664 | 0.4097 |
| 65 to 69 | 0.5977 | 0.8570 | 0.12116 | 0.2960 | 0.4283 |
| 70 to 74 | 0.4805 | 0.8408 | 0.15922 | 0.3449 | 0.4868 |
| 75 to 79 | 0.3461 | 0.8225 | 0.20101 | 0.4239 | 0.5972 |
| 80+ | 0.1000 | 0.8030 | 0.36056 | 0.5041 | 0.7167 |

The proportion of individuals requiring hospitalisation and an ICU stay (conditional on hospitalisation) were taken from the work of Salje et al[17]. We then used these figures in conjunction with recently updated estimates of the age-specific COVID-19 Infection Fatality Ratio (IFR, taken from Brazeau et al[35]) and the proportion of total deaths in each age-group occurring in i) the ICU and ii) in ICU patients receiving advanced respiratory support (ARS, taken from Knock et al[11]). We then calculated the age-specific probability of dying given a particular disease severity to match the IFR and setting-specific death proportions as follows:

$$CFR_{Hosp,a} = \frac{IFR_a}{Prob\ Hosp_a}$$

which gives us the overall Case Fatality Ratio in the hospital for age-group $a$, where $Prob\ Hosp_a$ is the probability of requiring hospital admission for age-group $a$. The $CFR$ for the ICU is then:

$$CFR_{ICU,a} = \frac{CFR_{Hosp,a} * PropDeath_{ICU,a}}{Prob\ ICU_a}$$

where $PropDeath_{ICU,a}$ is the proportion of hospital deaths in age-group $a$ occurring in the ICU, and $Prob\ ICU_a$ is the probability of requiring ICU admission for age-group $a$, conditional on hospitalisation. In turn:

$$CFR_{Crit,a} = \frac{CFR_{ICU,a} * PropDeath_{ARS\&ICU,a}}{Prob\ ARS_a}$$

is the CFR for Critical patients, where $PropDeath_{ARS\&ICU,a}$ is the age-specific proportion of ICU deaths occurring in patients receiving advanced respiratory support and $Prob\ ARS_a$ is the age-specific probability of receiving advanced respiratory support conditional on ICU admission. In turn, the CFR for Severe patients (representing all other patients in the ICU) is as follows:

$$CFR_{Sev,a} = \frac{CFR_{ICU,a} - (CFR_{Crit,a} * Prob\ ARS_a)}{(1 - Prob\ ARS_a)}$$

and finally, the CFR for moderately ill patients (those requiring oxygen but not a stay in the ICU) is calculated as:

$$CFR_{Mod,a} = \frac{CFR_{Hosp,a} - (CFR_{ICU,a} * Prob\ ICU_a)}{(1 - Prob\ ICU_a)}$$

the exact values for each of these parameters are available in Table S3.

**Table S6:** Prior Distributions for Fitted Model Parameters

| **Parameter** | **Prior** | **Description** |
|---|---|---|
| R0 | Normal | Mean = 3, standard deviation = 1, truncated such that R0 (i.e. the reproduction number in the absence of any non-pharmaceutical interventions) cannot be smaller than 1.6 and cannot be larger than 5.6. |
| Start Date | Uniform | Uniform in the range of 10-60 days prior to the first reported COVID-19 death. |
| $M\alpha$ | Normal | Mean = 0, standard deviation = 3, truncated such that the value cannot be larger than 10 or smaller than -10. |
| $M\omega$ | Uniform | Uniform in the range 0-1. |
| $\rho i$ | Normal | Mean = 0, standard deviation = 0.2, truncated such that the value cannot be larger than 5 or smaller than -5. |

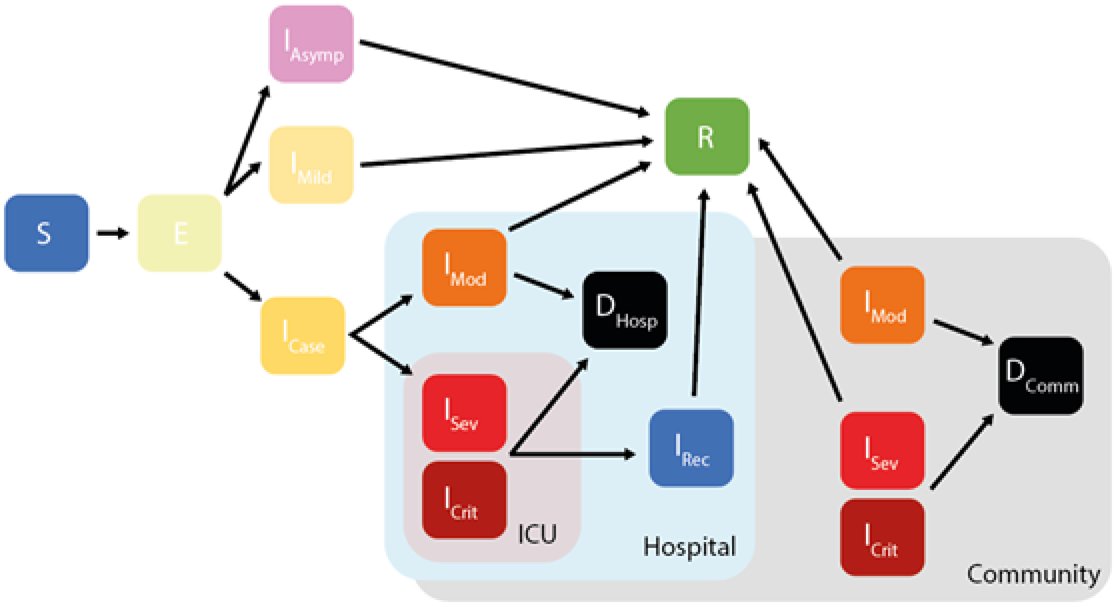


**Figure S1: Age-structured model of SARS-CoV-2 transmission explicitly incorporating disease severity, healthcare capacity and passage through different healthcare levels.** S = Susceptible, E = Exposed (Latent Infection), $I_{Asymp}$ = Asymptomatic Infection, $I_{Mild}$ = Mild Infections – neither Asymptomatic nor Mild infections require hospitalisation. $I_{Case}$ = Infections requiring hospitalisation (but who have not yet deteriorated to the point where they should be hospitalised). $I_{Mod-}$ = Infections with Moderate Disease (requiring a general hospital bed, minimal or low flow oxygen supplementation and associated care), $I_{Sev}$ = Infections with Severe Disease (requiring an ICU bed, high flow oxygen and associated care) and $I_{Crit}$ = Infections with Critical Disease (requiring an ICU bed, high flow oxygen, advanced respiratory support such as mechanical or non-invasive ventilation, and associated care). $I_{Rec}$ = Hospitalised Infections (requiring a general hospital bed after recovering from an ICU stay). R = Recovered and D = Dead. Pale blue box indicates compartments related to patients who, due to disease severity, require hospitalisation and are successfully hospitalised. Pale grey box indicates compartments related to patients who require hospitalisation due to disease severity, but who are not able to be hospitalised (due to lack of beds or reduced ability to access healthcare). For those who are successfully hospitalised, individuals pass through further decision points to determine the exact extent of clinical care they receive (determined by the availability of different healthcare components, see Figure S2 for more details). $D_{Hosp}$ = those who die in a healthcare setting (i.e. are successfully hospitalised) whilst $D_{Comm}$ = those who die in the community (i.e. are not successfully hospitalised).

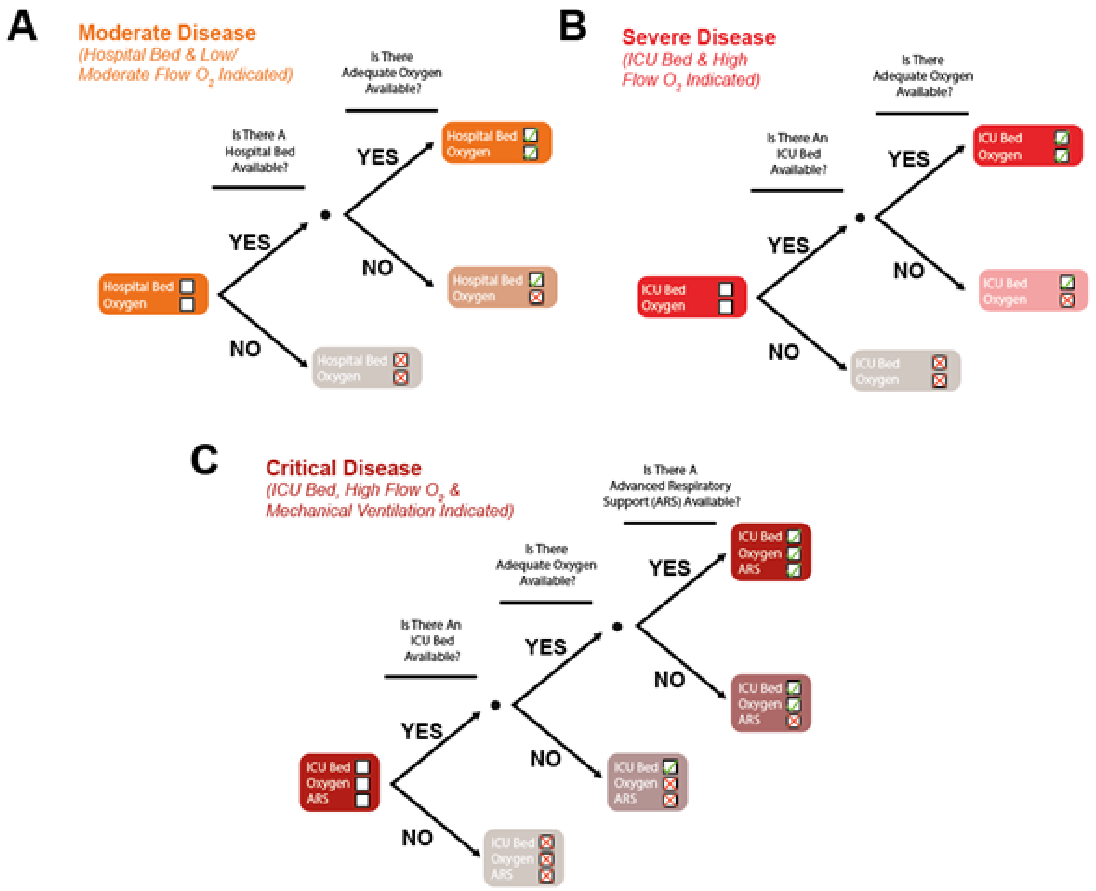


**Figure S2: Decision trees dictating healthcare individuals receive, for each of the disease severity categories modelled within the framework.** Within the framework utilised here, disease severity dictates what clinical care an individual requires. Healthcare capacity and availability dictates what clinical care an individual receives (with associated excess mortality when the clinical needs of an individual are not met). The modelling framework utilised here explicitly incorporates healthcare capacity and dynamically tracks the availability of several key healthcare components relevant to COVID-19 treatment. Specifically, it tracks the availability of hospital and ICU beds (and the associated general supportive care that being hospitalised would include), as well as oxygen (both low/moderate and high flow) and advanced respiratory support (either non-invasive machinces such as CPAP or biPAP as well as mechanical ventilators). Depending on their disease severity, individuals are cascaded through a series of decision trees assessing the availability of different healthcare components and receive that particular aspect of care if it is available. Conditional delivery of healthcare components is modelled such that, for example, receiving mechanical ventilation is dependent on mechanical ventilators being available, but also dependent on the availability of oxygen and an ICU bed. Decision tree pathway for (A) individuals with Moderate disease, which first assesses whether there is a general hospital bed available, and then, if there is, whether adequate oxygen is available, (B) individuals with Severe disease, which sequentially assesses whether there is an ICU bed high flow oxygen available for the individual. And (C) for individuals with Critical diseases, which assesses the availability of an ICU bed, high flow oxygen and of advanced respiratory support devices.

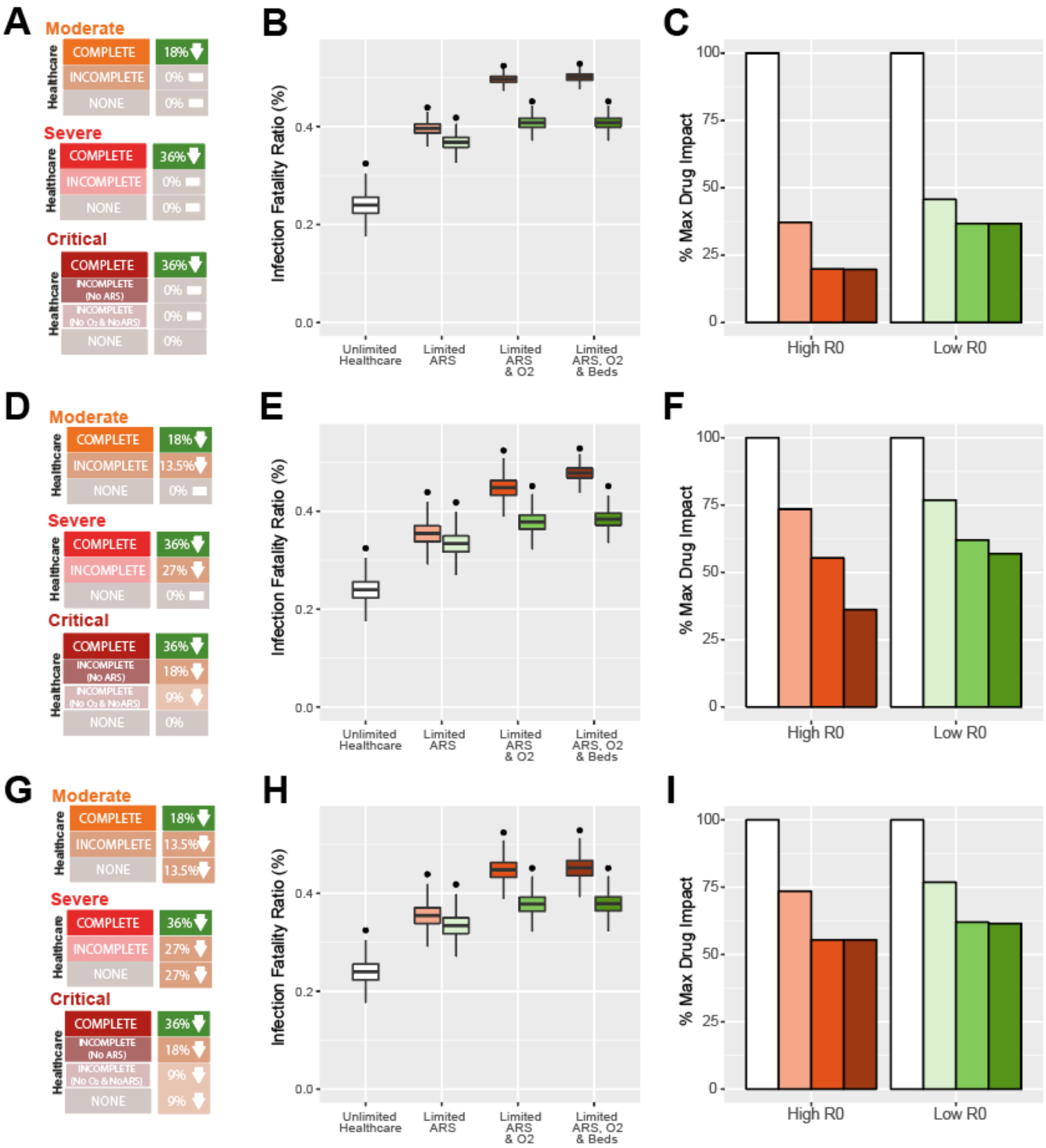


**Figure S3: The effect of varying assumptions surrounding dexamethasone's impact on COVID-19 mortality in the absence of other healthcare.** In addition to the results presented in the main text (Figure 2; which assumes full impact in fully-treated patients and limited impact in untreated/incompletely treated patients), we also explored a variety of different assumptions surrounding dexamethasone's impact. These range from assuming impact in fully treated patients only **(A)**, through to more optimistic levels of impact in incompletely treated patients **(D)** and additionally assuming some level of impact in individuals not hospitalised **(G)**. For each of these different scenarios, we present the infection fatality ratio (**B, E & H)** both with dexamethasone available (boxplots) and without (black dots), varying the availability of different healthcare components. We also present the percentage of dexamethasone's potential impact that is achieved under each of these scenarios **(C, F & I)**.

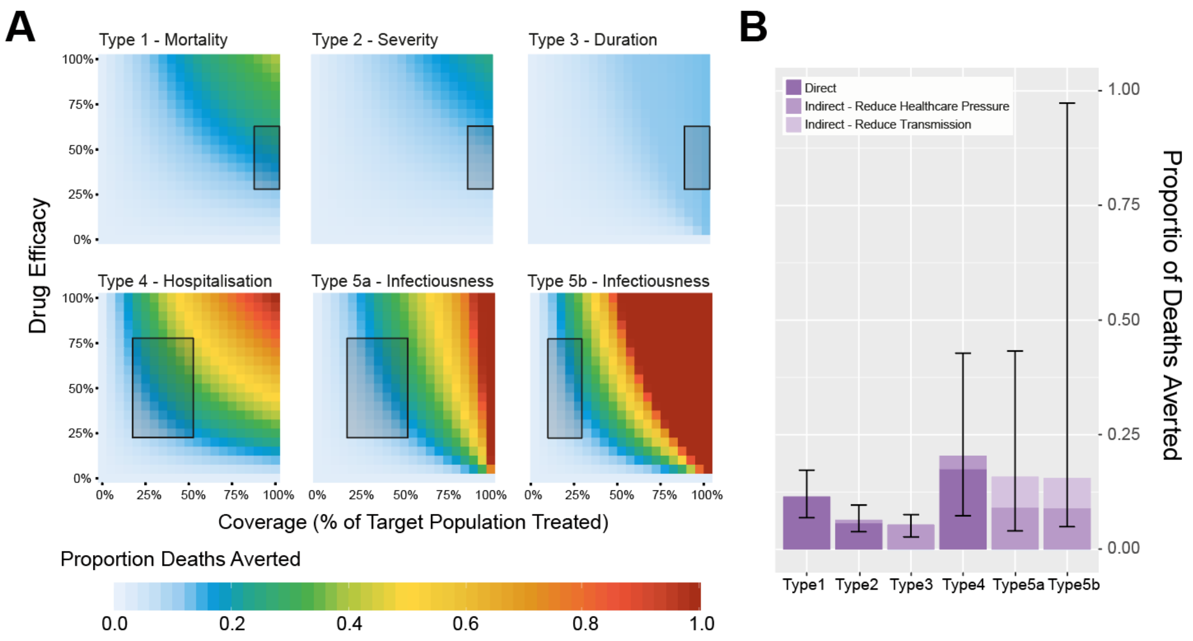


**Figure S4: Impact of different therapeutic product effects on COVID-19 disease burden, for a high R scenario. (A)** For an epidemic with an R of 2, the proportion of COVID-19 deaths averted as a function of therapeutic efficacy and therapeutic coverage, for 6 different types of potential effects (Table 1). These include reducing COVID-19 disease mortality (Type 1); preventing deterioration and worsening of disease in hospitalised patients (Type 2); reducing duration of hospitalisation (Type 3); preventing hospitalisation due to COVID-19 (Type 4) and reducing duration of infectiousness (Types 5a and Type 5b). Inset boxes indicate the range of plausible values of coverage used to generate the estimates in **(B)**. **(B)** Disaggregation of therapeutic effect type impact by whether this is direct or indirect. Bars are coloured according to the type of impact (direct reduction in mortality, indirect reduction in mortality due to reduced pressure on healthcare or indirect reduction in mortality due to reductions in community transmission), with error bars indicating the maximum and minimum proportion of deaths averted under the range of coverage and effectiveness values considered for each effect type (indicated by the boxes in **(A)** and **Table 1**).
